# Protocol for ACCESS D: a mixed-methods feasibility study of a community-based model to improve equity and efficiency in dementia research participation

**DOI:** 10.64898/2026.02.04.26344869

**Authors:** Patricia Fuller, Andy Claxton, Helen Pocock, Samantha Williams, Nicola Claxton, Amanda Wollam, Daniel Blackburn, George Devitt, Sarah Fearn, Christopher Kipps

## Abstract

**Background:** Despite national efforts to improve research inclusion, people from underserved communities remain underrepresented in dementia trials. Barriers occur at the point of initial engagement and also within the participation pathway itself, as the structure and burden of early screening procedures can discourage continuation. ACCESS D (Advancing Community Collaboration and Engagement Strategies in Dementia) aims to address these challenges by testing a community-based model that combines co-produced engagement events, low-burden research activities, and real-time support from the South Central Ambulance Service (SCAS), a trusted, community-visible healthcare workforce.

**Methods and analysis:** ACCESS D is a 12-month mixed-methods feasibility study recruiting 100 adults aged 50–90 years with either (i) a diagnosis of mild cognitive impairment or dementia or (ii) a self- or proxy-reported memory concern affecting daily life. The study will deliver 12–18 co-produced community outreach events in non-clinical settings, supported by SCAS research paramedics and nurses. Following consent, participants will complete a core questionnaire and may optionally take part in one or more low-burden research activities designed to provide supported, first-hand experience of dementia research. Feasibility outcomes, including pathway progression and opt-in to future dementia research contact, will be descriptively summarised and stratified using NIHR INCLUDE-aligned underserved characteristics. Qualitative interviews and focus groups with participants and staff will examine acceptability, perceived value, barriers and enablers, and implementation learning, analysed using thematic analysis and integrated with quantitative findings.

**Ethics and dissemination:** The study has received a favourable opinion from the Southwest Frenchay Research Ethics Committee and Health Research Authority approval (IRAS 361074). Findings will be disseminated via peer-reviewed publications, conference presentations and co-produced lay outputs for community partners and participants. These outputs will be accompanied by an implementation toolkit for research teams and a visual summary for potential participants.

**Strengths and limitations of this study:**

- This study tests a community based, co-produced delivery model that embeds real research participation within outreach, addressing both awareness and structural barriers to dementia research participation.
- Outreach is delivered by a trusted, community visible healthcare workforce, enabling real time support and reducing psychological, practical and digital barriers for underserved groups.
- The mixed methods feasibility design combines quantitative indicators of reach, progression and resource use with qualitative insights into participant and staff experience, generating actionable learning for scale up.
- Outcomes are explicitly equity-stratified using NIHR INCLUDE aligned characteristics, allowing early assessment of differential reach and engagement across underserved populations.
- As a single region feasibility study, findings may have limited immediate generalisability. However, the study is designed to generate transferable implementation insights and inform a future multi-site evaluation.

## Background and rationale

Dementia affects over 980,000 people in the UK and 57 million worldwide, with numbers expected to double by 2050.^1^ It disproportionately affects older adults, but its impact is felt much more widely, among families, caregivers, communities and healthcare systems. The economic cost is immense; it costs the UK over £42 billion annually^2^ and global costs are predicted to rise from $1.3 trillion to $2.8 trillion by 2030.^3^

Research and clinical trials are essential to advance diagnosis, care and treatment, yet participation remains low and non-diverse. Barriers include mistrust, low awareness, stigma, fear of diagnosis, and limited access to research opportunities, particularly in underserved populations where dementia risk is often higher.^4^ Traditional research models, largely based in hospitals or academic centres, increasingly focus on early-stage disease, when symptoms are subtle or mistaken for normal ageing. And even when outreach raises awareness, many people disengage due to the complexity, timing, or intrusiveness of early screening. ^5-8^

As a result, dementia research participants tend to be predominantly White, highly educated, and already positive about research.^5,9^ This creates a cycle of exclusion. Communities most affected by dementia miss out; not only on the chance to influence future care and access new treatments, but also on the personal benefits of participation.^10,11^

These inclusion gaps reduce the efficiency and generalisability of dementia research.^12 13^ For people affected by dementia, this contributes to slower innovation, inequitable access to advances, and interventions that may not perform optimally across diverse populations, undermining the effectiveness of dementia care and the overall resilience of healthcare systems. Moreover, as dementia research increasingly prioritises early-stage and prevention trials, understanding how people enter, experience and progress through research participation pathways has become critical, aligning with key priorities of the NIHR INCLUDE Framework, the Dementia Mission, and the UK Life Sciences Vision.

ACCESS D addresses these longstanding structural and equity barriers by developing and testing a participant-centred, community-based model. Rather than expecting people to navigate to research opportunities within clinical and academic settings, ACCESS D brings dementia research directly to diverse community environments, making research more accessible and less intimidating.

Previous studies have shown that community-led strategies can increase participation by building trust, using culturally relevant messaging, and involving local champions.^12-17^ However, persistent gaps remain, especially among people with lower income, lower educational attainment, or limited understanding of research.^18-20^ Misunderstandings between research and clinical care, and uncertainty about the benefits of participation, remain common.^21-23^ As dementia research increasingly explores risk disclosure and prevention, transparent and accessible communication is essential to support informed decision-making.^24,25^

ACCESS D responds to these challenges by co-developing inclusive community events with people affected by dementia, public contributors (PPIEP), academic researchers, and the South Central Ambulance Service (SCAS) research team. Events are delivered in familiar locations, including areas of increased socio-economic deprivation and communities historically underrepresented in dementia research.

A persistent structural gap exists between raising awareness of research and enabling meaningful participation. ACCESS D seeks to bridge this gap by embedding opportunities for research participation within community engagement events, supported in real time by the SCAS research team. This approach allows participants to experience what dementia research involves in a supported, low-pressure environment, helping to reduce both psychological and structural barriers to participation and create more equitable entry points into dementia research pathways.

High screen failure and early drop-out are well recognised in dementia research, particularly in early-phase and prevention trials. These inefficiencies can be frustrating for participants who have invested time and hope and are costly for research teams.^26^ ACCESS D examines where interest develops or stalls along the participation pathway and whether real-time support and embedded research activities can help align expectations, improve acceptability and sustain engagement. By engaging participants earlier and in more accessible settings, the study aims to generate feasibility data to inform strategies for reducing screen failure and early dropout and supporting pathway progression.

ACCESS D is designed to generate learning in both directions. For community members, it offers a safe, non-clinical space to ask questions, experience research first-hand, and share views on dementia, perceived risk, and study design. For the research system, it generates structured insight into who is reached, what support is needed, which elements are acceptable and where barriers persist across the participation pathway. In parallel, ACCESS D will generate feasibility and process data on resource use and deliverability of the model. These data are intended to inform future programme-level funding, guide decisions about wider implementation, and support adaptation of the model to other conditions and settings.

Dementia is used as a test case to evaluate the broader ACCESS framework: a flexible, community-first approach designed to improve equity and efficiency in research participation across conditions. The framework is underpinned by three interdependent principles:

1. Co-production with communities to ensure cultural relevance and legitimacy
2. Delivery by trusted messengers to enhance credibility and approachability
3. Low-burden, supported research participation to reduce barriers and build confidence

By explicitly testing these principles in practice, ACCESS D aims to contribute transferable evidence to support decentralised, inclusive research delivery aligned with national priorities.

## Study Aim and Objectives

### Aim

To evaluate whether community-based outreach and support can make access to dementia research opportunities more inclusive and more efficient.

### Objectives

- Co-develop and pilot ways of engaging underserved communities that reduce barriers and help people consider taking part in dementia research.
- Explore the feasibility and acceptability of low-burden research activities, such as questionnaires, interviews, digital tests, and finger-prick samples, and their impact on confidence and research readiness.
- Assess early indicators of reach, pathway progression, and scalability to inform future strategies for more inclusive and efficient recruitment.

## Methods and Analysis

### Study design

ACCESS D is a 12-month mixed-methods feasibility study designed to test the feasibility, acceptability and implementation of a community-based model which aims to improve the equity and efficiency of dementia research participation.

In ACCESS D, “efficiency” refers to early pathway progression and resource use. It will be assessed using stage-specific conversion rates (exposure/interest → consent → completion of ≥1 activity → opt-in to future contact) and delivery resource metrics (staff time and direct delivery costs per consented participant and per opt-in).

The study is structured around four linked work packages (WP; see Fig 1):

**Figure 1.**
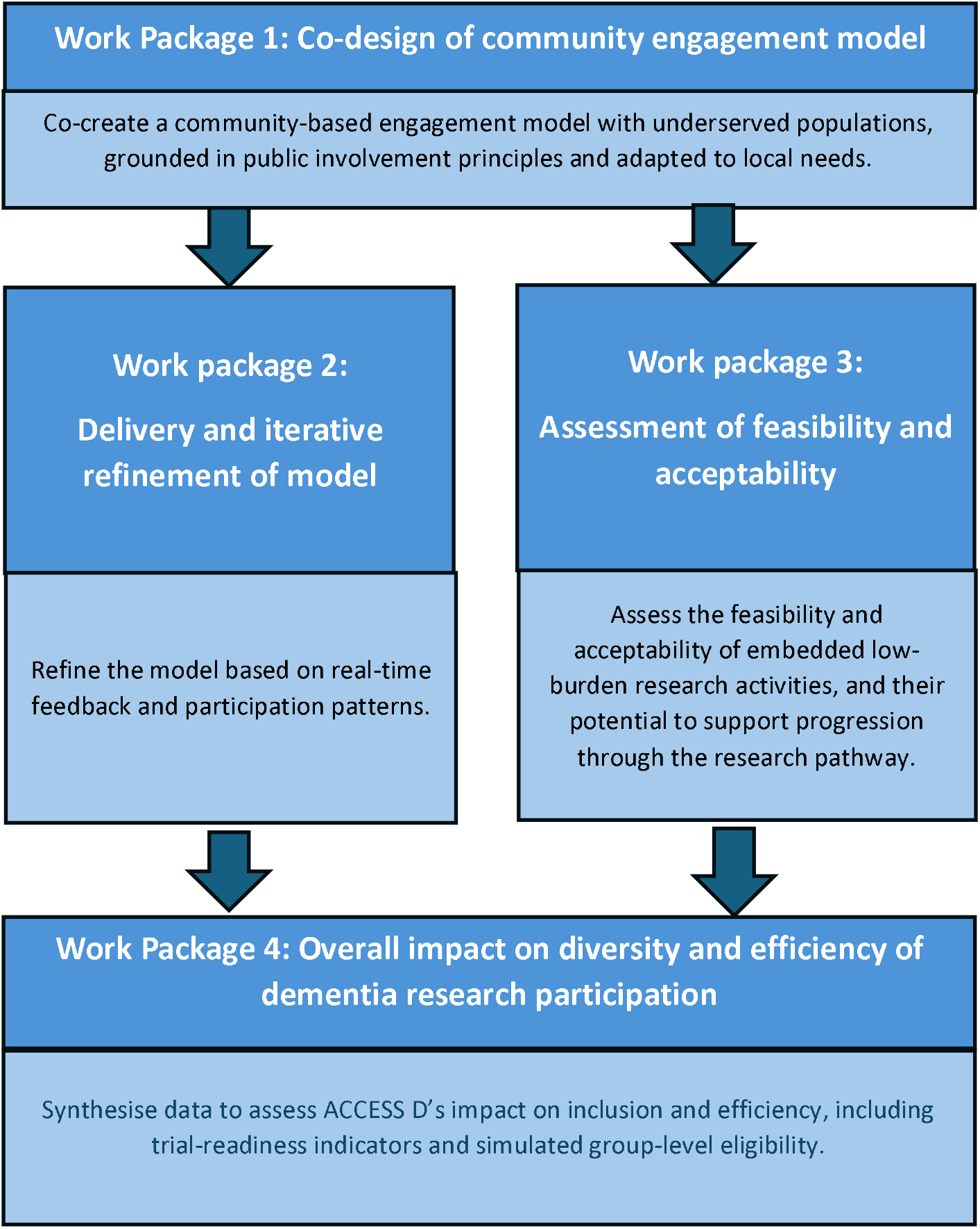
Conceptual overview of the ACCESS D work package structure

#### WP1: *Co-produce a community-based engagement model*

This work package involves co-developing an outreach model with people from underserved communities, public contributors, the SCAS research team, and academic researchers. A minimum of three co-production workshops will be held in accessible, non-clinical community settings. These sessions will explore culturally relevant messaging, inclusive event design, trust-building strategies, and barriers to research participation. Live illustration will be used to support accessibility. Outputs will include a co-produced event format, a visual summary for participants and a delivery toolkit for delivery teams.

#### WP2: *Implement and refine the outreach model*

A minimum of 12–18 dementia-focused community outreach events will be delivered over six months across diverse locations (e.g. faith centres, public events, community groups). Events will be led by the SCAS research team and offer attendees opportunities to learn about dementia, ask questions, and optionally take part in real research activities (e.g. questionnaires, testing a new digital cognitive assessment (<u>CognoMemory</u>), fingerprick sampling for biomarkers). Real-time feedback will be captured via debriefs and implementation logs to support ongoing refinement of the model.

#### WP3: *Evaluate feasibility, acceptability and value*

This WP will assess the feasibility, acceptability, and perceived value of offering embedded research activities in community settings. Data sources include post-event questionnaires (Likert and free-text), interviews and focus groups with participants, and reflective interviews with SCAS staff. Findings will explore motivators, barriers, and how these activities shape understanding and confidence around research participation and sustained engagement.

#### WP4: *Evaluate progression and diversity across the participation pathway*

The final work package will synthesise data to assess the model’s impact on both diversity and efficiency. This includes analysis of pathway progression (e.g. consent and completion rates), dropout points, and simulated trial eligibility. Outcomes will inform the design of more inclusive dementia research pipelines and provide a foundation for future scale-up.

### Setting

ACCESS D will be delivered across non-clinical community venues in Southampton and surrounding areas, selected co-productively with public contributors, community partners, research paramedics and nurses and the academic team to maximise accessibility, cultural relevance and inclusion. Likely settings include community centres, places of worship, libraries, public events and other community-facing locations. Venue suitability (including privacy and the feasibility of on-site research activities) will be assessed in advance through risk assessment led by SCAS. Where required, a SCAS research ambulance will provide a mobile private space for consenting and optional finger-prick sampling.

### Patient and public involvement and engagement

ACCESS D has been developed with public contributors. Deliberative dialogue sessions highlighted limited public understanding of what research involves and SCAS community workshops revealed strong support for community-based, face-to-face opportunities to ask questions and experience research in a supported, low-pressure setting. Public contributors reviewed and informed the study rationale, participant information sheets, consent forms and questionnaire, and will work with the research team to interpret and disseminate findings, including co-produced lay outputs, presentations and peer reviewed publications.

### Recruitment pathways

To maximise reach and enable descriptive comparison of engagement profiles, ACCESS D will recruit participants via four pre-specified pathways (see Fig 2):

1. **Community events:** in-person recruitment in community venues led by the SCAS research team, with optional on-site research participation.
2. **NHS memory clinics:** clinical teams introduce the study to potentially eligible patients, for example those who although hesitant to enrol on a trial, would be keen to know more about research first before committing; individuals can enrol online and/or attend a community event.
3. **Research registries:** recruitment via existing platforms (e.g., Join Dementia Research).
4. **Online recruitment:** adverts and leaflets linking to online study information and consent.

**Figure 2.**
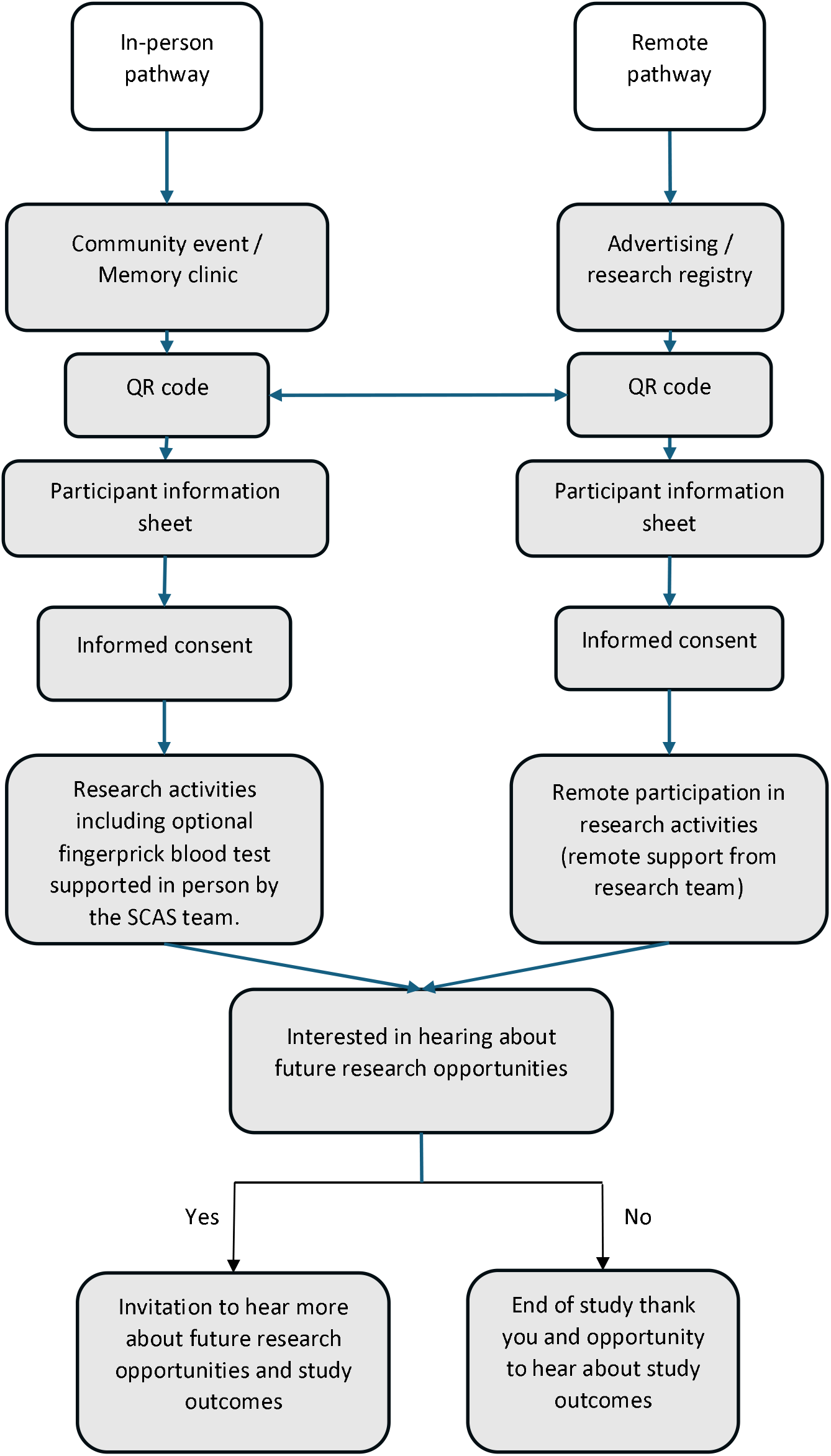
ACCESS D participation pathways

Participants may move between pathways. For example, someone who registers online may choose to attend a community event, or a memory clinic patient may opt to complete activities remotely. These transitions will be recorded and disaggregated in analysis. The target of 100 participants is cumulative across all routes.

### Outreach events and implementation

A minimum of 12–18 outreach events will be delivered over approximately six months. Events will vary in duration (approximately 2–6 hours) depending on venue type and footfall (e.g., organised group sessions versus public pop-ups). At events, attendees can:

- learn about dementia and dementia research using NHS-approved materials,
- speak with the SCAS research team in an informal setting, and
- optionally consent and take part in study activities with real-time support.

Participation in research activities is voluntary; attendees may engage with events without taking part in any research. The delivery format will be iteratively refined using implementation logs, event feedback and debriefs to identify barriers, improve acceptability and optimise accessibility.

### Participants

#### Eligibility Criteria

Participants will be enrolled into two broad groups:

- **Group A:** Those with a diagnosis of mild cognitive impairment (MCI) or dementia. This group includes individuals with established clinical diagnoses, to understand engagement in populations currently accessing clinical care
- **Group B:** Those with memory concerns but no formal diagnosis.

#### Inclusion Criteria

- Aged 50–90 years (inclusive).
- Group A: clinical diagnosis of MCI or dementia.
- Group B: self- or proxy-reported memory concerns perceived to affect daily life.

#### Exclusion Criteria

- Lack of capacity to provide informed consent at enrolment.
- Any medical, psychiatric or behavioural condition which, in the opinion of research staff, would make participation unsafe.

### Capacity and informed consent

All participants must have capacity to provide informed consent, assessed in accordance with the Mental Capacity Act (2005). Capacity will be assumed unless there is evidence to the contrary. Trained research staff will ensure participants can understand, retain and weigh the study information and communicate a voluntary decision.

Consent will be given before any study activities. Participants may consent in person (paper or electronic) at outreach events, or remotely via a secure online platform accessed through a QR code. Individuals can take information away and decide in their own time, and can ask questions at any point. Participants may withdraw at any time without giving a reason; where feasible, anonymised reasons for withdrawal will be recorded to inform feasibility and improvement.

### Study procedures and data collection

Study procedures and data collection are summarised in Table 2.

**Table 2.** Summary of ACCESS D study activities, settings and timing.

| Activity | Setting | Timing | Data Captured |
| --- | --- | --- | --- |
| Participant information sheet and informed consent | 1. Community event<br>2. Memory clinic<br>3. Registry<br>4. Fully remote | Enrolment | Paper or electronic consent |
| Core questionnaire | Event tablet or personal device | Post consent | Primary outcome, including demographics, ECog 12/ experience/acceptability/views on memory, dementia risk and research design. |
| Optional digital memory test | Event tablet or personal device | Post consent | Summary outputs from CognoMemory |
| Finger-prick sample | Community events only | Post consent | Exploratory biomarkers including for example amyloid proteins |
| Focus group / interview | Remote or in-person | Post participation | Purposive sampling including underserved status and those who decline research activities |
| 6-month follow up email | Remote via email | Approx 6 months post participation | Self-report of subsequent research participation |

### Overview of study activities

Following informed consent, participants will be assigned a unique study ID and offered:

- a core study questionnaire (all participants), and
- optional research participation opportunities (participants may choose any, all or none).

### Core activity: Questionnaire

All participants will be offered a mixed-format questionnaire (Likert and free-text) available in paper or digital format, with staff support as needed. The questionnaire covers:

- experience and acceptability of the outreach event and any research activities completed
- perceived barriers and enablers to dementia research participation
- confidence and willingness to take part in future research
- views on dementia and research design
- self-reported memory concerns/diagnoses
- representation variables (age, gender, ethnicity, education, postcode to derive Index of Multiple Deprivation)
- the Everyday Cognition Questionnaire (ECog-12)
- consent preferences for future research contact and optional follow-up.

### Optional research participation opportunities

1. **Digital memory assessment (CognoMemory):** a web-based, speech-driven memory assessment completed either at events (provided device or participant’s own) or remotely on a participant’s device, taking approximately 10 minutes. A separate digital consent process is embedded within the CognoMemory platform under its own ethical approval.
2. **Finger-prick blood sampling (events only):** participants attending in person may opt to provide a finger-prick blood sample for dementia biomarkers and collected by the SCAS research team in an appropriate private setting.
3. **Qualitative interview or focus group:** a purposive subsample of participants will be invited to take part in an interview (up to 45 minutes) or focus group (up to 60 minutes), conducted in person, online or by telephone. Sessions will be audio-recorded with consent, transcribed and analysed thematically. Sampling will include participants from underserved groups and those who declined optional activities.
4. **Six-month follow-up:** participants may opt to receive a brief follow-up email approximately six months after participation asking whether they have taken part in other dementia research since ACCESS D.

### Role of illustrative research activities

Research activities offered within ACCESS D are not being formally evaluated for clinical validity. Instead, they serve two distinct purposes: (1) as low-burden examples of typical dementia research procedures, enabling participants to experience deconstructed trial components (e.g., interviews, questionnaires, digital cognitive tests, finger-prick sampling) in a safe, supported setting; and (2) to generate feasibility data on delivery, acceptability, and uptake in community settings, and to explore whether the ACCESS D model can increase the diversity and efficiency of research participation.

### Staff interviews and implementation data

SCAS research team staff involved in delivery will contribute implementation insights through debrief discussions, event evaluation logs and semi-structured interviews. Topics will include feasibility of delivery, workload, barriers and enablers, participant support needs, and recommendations for optimisation and scale-up.

### Outcomes

#### Primary outcome: progression to future research contact

The primary outcome is the proportion of consented participants who opt in to be contacted about future dementia research opportunities.

- **Numerator:** number of participants providing consent for future research contact.
- **Denominator:** all consented participants.

This outcome was selected as a key indicator of feasibility because it reflects a key transition from initial engagement to potential longer-term involvement in research without relying on the availability of a specific downstream trial. It captures participants’ willingness to remain connected to research and helps assess whether the ACCESS D model can support sustained research readiness among diverse and underserved populations.

#### Co-primary outcome: equity-stratified primary outcome

The primary outcome will be stratified by underserved status (NIHR INCLUDE–aligned criteria) and by recruitment pathway to assess whether ACCESS D reaches and engages groups historically underrepresented in dementia research.

For analysis, participants will be classified as “underserved” if they meet ≥1 of the following pre-specified criteria: (i) Index of Multiple Deprivation (IMD) deciles 1–3 derived from participant home postcode (socioeconomic deprivation); (ii) minority ethnic background (UK standard ethnicity categories; self-report); (iii) low digital confidence (self-report), (iv) low educational attainment (self-report). In addition, we will derive an underserved score (0–4) as the count of criteria met to support descriptive summaries of intersectionality.

### Secondary outcomes

Secondary outcomes will be analysed descriptively, and will be stratified by underserved group status where numbers permit.

#### Process efficiency

- Activity uptake: proportion of consented participants completing ≥1 study activity (questionnaire and an optional activity).
- Delivery fidelity: proportion of planned outreach events delivered.
- Event yield: number of participants consented per outreach event and number completing ≥1 activity per event.

#### Staff experience

- Staff acceptability: proportion reporting a positive post-event experience (≥7/10).
- Staff time: staff hours per event, by role.
- Qualitative staff experience: themes from staff interviews, debriefs and delivery logs.

#### Participant acceptability

- Overall experience: proportion rating experience as positive (≥7/10).
- Recommendation: proportion willing to recommend participation to others.
- Support needs: proportion requiring assistance to complete any activity.
- Qualitative participant experience: themes relating to trust, burden, clarity, perceived value and barriers.

### Pathway efficiency (within-participant progression)

- Stage-specific conversion rates across the participation pathway (exposure/interest → consent → completion of ≥1 activity → opt-in to future contact → optional 6-month follow-up response), summarised overall and stratified by recruitment pathway.

### Recruitment pathway yield (exploratory)

- Exploratory analysis of opt-in rates by recruitment pathway to provide indicative signals of yield. Denominators are defined as:
  - **Community events:** number of individuals who engage with the outreach team and accept study information (leaflet/QR card) as recorded in event logs.
  - **NHS memory clinics:** number of study leaflets distributed (clinic log).
  - **Research registries (e.g**., **JDR):** number of invitations issued (and, where available, number opened/clicked) as provided by the registry platform.
  - **Online recruitment:** number of unique study-page visits (and, where available, impressions and click-throughs) from advertisement analytics.

### Trial eligibility simulation (exploratory)

- Group-level simulation of eligibility for exemplar dementia trials using questionnaire data, demographics, and optional activity data (further described in Data Analysis).

### Longer-term engagement (exploratory)

- Response to optional 6-month follow-up and self-reported participation in other dementia research since ACCESS D.

### Resource use and scalability (exploratory; at event and project level)

- Staff hours (by role)
- Direct delivery cost
- Cost per consented participant
- Cost per opt-in to future research contact

### Sample size

This is an exploratory feasibility study; therefore, no formal sample size calculation has been conducted. A pragmatic target of 100 participants typical of feasibility studies has been set to assess feasibility and acceptability in diverse community settings and to provide initial descriptive estimates of engagement and progression, including stratified summaries where subgroup sizes permit. For qualitative components, up to 20 participants and approximately 5 SCAS staff will be purposively sampled to ensure diversity and analytic adequacy.

## Data analysis

### Quantitative analysis

Quantitative analysis will be descriptive, with no hypothesis testing. Analyses will be conducted using STATA (or equivalent). Categorical variables will be summarised using frequencies and proportions; continuous variables using means and standard deviations or medians and interquartile ranges, as appropriate. Where subgroup numbers permit, outcomes will be stratified by underserved group status and recruitment pathway. Missing data will not be imputed; patterns of missingness (e.g., incomplete surveys or non-completion of optional activities) will be described to inform future studies.

### Exploratory group-level simulation of trial eligibility

In the absence of an actively recruiting dementia trial during the study period, data collected within ACCESS D (e.g. age, ECog-12, and optional measures such as CognoMemory summaries and biomarkers) will be used in an exploratory, group-level simulation. This simulation will estimate the proportion of participants who would theoretically meet pre-specified exemplar dementia trial eligibility criteria, selected to reflect common inclusion and exclusion features of early-phase dementia trials, and to characterise patterns of potential exclusion.

This analysis is intended solely to inform understanding of eligibility constraints at a population level. It will not be used to make individual eligibility determinations, and no individual results will be returned to participants.

### Qualitative analysis

Free-text survey responses and interview/focus group transcripts of participants and staff will be analysed using inductive thematic analysis following Braun and Clarke’s six-phase approach. NVivo and/or Excel will support data management. Interpretation will be guided by relevant theoretical lenses, used to contextualise findings rather than as prescriptive coding frameworks, including Normalisation Process Theory and the Theoretical Framework of Acceptability, to explain acceptability and implementation across contexts (i.e., what works, for whom, and in what settings).

### Mixed-methods integration

A convergent mixed-methods design will be used. Quantitative and qualitative findings will be analysed separately and then integrated at interpretation through triangulation to develop overall conclusions about feasibility, acceptability, equity and scalability of the ACCESS D model. Integration will follow a triangulation protocol with comparison matrix.

### Study management, governance, and ethics

ACCESS D will be overseen by a Study Management Group (SMG) comprising the Chief Investigator, the study coordinator and lead applicant, South Central Ambulance Service (SCAS) research paramedic delivery lead, and Patient and Public Involvement and Engagement (PPIEP) contributors. The SMG will oversee study delivery, recruitment and equity monitoring, participant safety, data quality, and documented iterative refinements to the delivery model.

The study has received a favourable opinion and approval from the Southwest Frenchay Research Ethics Committee and the Health Research Authority. Any protocol amendments will be submitted for REC/HRA approval as required prior to implementation.

ACCESS D will comply with the UK General Data Protection Regulation (UK GDPR), the Data Protection Act 2018, and sponsor and university information governance policies. All participants will be assigned a unique study identifier. Research datasets will be pseudonymised, with identifiable information stored separately and accessed only by authorised study personnel.

Consent and questionnaire data will be captured using secure, institutionally approved systems (e.g. REDCap or Qualtrics). Access will be role-restricted, and analysis datasets will contain no direct identifiers. Data generated through the CognoMemory platform will be governed under separate ethical approval and consent processes; only minimised, pseudonymised summary outputs required for pre-specified exploratory analyses will be shared with the ACCESS D team under appropriate data sharing agreements.

Participants opting into finger-prick sampling will provide samples labelled with study ID only, processed in accordance with sponsor and laboratory governance requirements. Qualitative interviews and focus groups will be audio-recorded with consent, transcribed, de-identified and stored securely.

ACCESS D is designed to minimise risk. Optional study activities are research-only and non-diagnostic, and no individual results will be returned to participants, as detailed in the participant information sheets. Staff will follow a distress protocol to identify and respond to emotional discomfort, including pausing activities, reaffirming voluntariness and signposting to appropriate support. Participants may withdraw at any time without consequence; unless otherwise requested, data collected up to withdrawal will be retained in pseudonymised form.

## Discussion

ACCESS D is a mixed-methods feasibility study designed to address persistent inequities and inefficiencies in dementia research participation. It combines co-produced community outreach, delivery by a trusted healthcare workforce, and real-time, supported opportunities to experience low-burden research activities typical of dementia studies and trials. The goal is to make research more accessible, better understood, and more inclusive, particularly for people from underserved communities and those not yet known to memory services.

The study will generate descriptive feasibility and process data on reach, progression, equity, and resource use, alongside qualitative insights into participant and staff experience to ensure acceptability. Findings will inform future programme-level applications and provide practical guidance for embedding community-based, equitable, and efficient approaches within dementia research infrastructure.

Following this feasibility phase, the next step is a multi-site implementation study to test transferability across regions, delivery contexts and conditions. Future work will also explore data linkage to routine health records and research registries to assess sustained engagement and longer-term outcomes.

Although ACCESS D focuses on dementia, its core principles of community co-production, delivery by trusted messengers, and low-burden, supported research activities are intentionally designed to be transferable to other long-term conditions, such as respiratory disease, where similar participation challenges exist. The model also contributes to national priorities set out in the NIHR INCLUDE Framework, the Dementia Mission, and the UK Life Sciences Vision, supporting more inclusive recruitment, minimising early attrition and screen failure, and enabling more efficient research delivery.

## Data availability statement

De-identified quantitative and qualitative data generated during the study may be available on reasonable request from the corresponding author, subject to appropriate governance approvals and data sharing agreements.

## Author contributions

PF: Conceptualisation, methodology, Investigation, funding acquisition, writing – original draft, writing – review and editing.

AC: Conceptualisation, methodology, Investigation, writing – review and editing

HP: Methodology, Investigation, writing – review and editing.

SW: Methodology, Investigation, writing – review and editing.

NC: Methodology, Investigation, writing – review and editing.

AW: Methodology, investigation, writing – review and editing.

DB: Methodology, investigation, writing – review and editing.

GD: Methodology, investigation, writing – review and editing.

SF: Methodology, investigation, writing – review and editing.

CK: Conceptualisation, methodology, writing – review and editing.

